# Associations between evening shift work, irregular sleep timing, and gestational diabetes in the Nulliparous Pregnancy Outcomes Study: Monitoring Mothers-to-be (nuMoM2b)

**DOI:** 10.1101/2022.05.23.22274967

**Authors:** Danielle A Wallace, Kathryn Reid, William A Grobman, Francesca L Facco, Robert M Silver, Grace W Pien, Judette Louis, Phyllis C Zee, Susan Redline, Tamar Sofer

## Abstract

**Study Objectives:** Shift work is a risk factor for cardiometabolic disease, possibly through effects on sleep-wake rhythms. We hypothesized that shift work during pregnancy is associated with increased odds of preeclampsia, preterm birth, and gestational diabetes mellitus (GDM), and that the association is mediated by irregular sleep timing.

**Methods:** The Nulliparous Pregnancy Outcomes Study: Monitoring Mothers-to-be (nuMoM2b) is a prospective cohort study (n=10,038) designed to investigate risk factors for adverse pregnancy outcomes. Medical outcomes were determined with medical record abstraction and/or questionnaires; sleep midpoint was measured in a subset of participants with ≥5-day wrist actigraphy (ActiWatch). We estimated the association of shift work during pregnancy with preeclampsia, preterm birth, and GDM using logistic regression, adjusted for adversity (a cumulative variable for poverty, education, health insurance, and partner status), smoking, self-reported race/ethnicity, and age. Finally, we performed an analysis to explore the extent to which to the association between shiftwork and GDM was mediated by variability in sleep timing.

**Results:** Evening shift work during pregnancy is associated with approximately 75% increased odds of developing GDM (adjusted OR=1.75, 95% CI:1.12-2.66); there was no association with preterm birth and no association with preeclampsia after adjustment. Pregnant evening shift workers were found to have approximately 45 minutes greater variability in sleep timing compared to day workers (p<0.005); mediation analysis estimated that 27% of the association between shift work and GDM was explained by sleep-timing variability.

**Conclusions:** Evening shift work was associated with GDM, and this relationship may be mediated by variability in sleep timing.

## INTRODUCTION

Shift work is a common occupational exposure, with approximately 15-20% of the U.S. workforce employed in some form of shift work outside of the working hours of 7AM-5PM(1). As the majority of pregnant people continue to work while pregnant(2), shift work may be a common exposure during pregnancy. Shift work may have adverse effects on pregnancy due to its impact on health outcomes such as diabetes(3,4), dyslipidemia(5), and other cardiometabolic disease (CMD)(6,7). In addition, the International Agency for Research on Cancer has categorized night shift work as a “probable” human carcinogen (Group 2A)(8).

The effects of shift work during pregnancy on health warrant further study. Shift work is proposed to adversely affect pregnancy outcomes via circadian misalignment and impaired sleep quality and quantity, which can cause endocrine disruption and alter metabolic and growth processes(9). However, the evidence to support an association between shift work and adverse pregnancy outcomes is mixed. A recent systematic review and meta-analysis of shift work during pregnancy and adverse outcomes concluded that both night and rotating shift work were associated with increased odds for preterm birth, but there was little evidence to support an association with preeclampsia(10). Other recent cohort studies reported a null association(11) and a positive association(12) between night shift work and preterm birth. Only one prior study has evaluated shift work and GDM(13), and reported no association.

By definition, shift workers have work schedules that differ from standard working hours. As a consequence, the timing of other daily activities, such as sleep, are also altered. As irregular sleep schedules or variability in sleep timing are also linked to increased risk of CMD(14–20), irregular sleep timing may be a dimension of shift work that contributes to these adverse health impacts. Sleep timing may also influence health during pregnancy. Self-reported later midpoint of sleep was previously found to be associated with preterm birth(21) and GDM(22), and objective (as measured by actigraphy) later sleep midpoint (>5 AM) was associated with GDM(23) in the Nulliparous Pregnancy Outcomes Study: Monitoring Mothers-to-be (nuMoM2b), a large prospective birth cohort study. Therefore, shift work may be a risk factor for preeclampsia, preterm birth, and/or GDM via irregular sleep schedules.

While both shift work and impaired sleep are associated with adverse pregnancy outcomes, no prior study has evaluated the role of whether objectively measured sleep timing mediates the relationship between shift work and pregnancy outcomes. Additionally, few studies have examined shift work during pregnancy or irregular sleep timing during pregnancy and GDM. Therefore, to address these gaps in knowledge, we analyzed data from the Nulliparous Pregnancy Outcomes Study: Monitoring Mothers-to-be (nuMoM2b) cohort. We hypothesized that shift work during pregnancy is associated with adverse outcomes (i.e., preeclampsia, preterm birth, and GDM) and explored whether associations are mediated by irregular sleep timing.

## METHODS

### Study population and cohort design

The Nulliparous Pregnancy Outcomes Study: Monitoring Mothers-to-be (nuMoM2b) is a prospective birth cohort study in the U.S. that enrolled 10,038 participants with viable, singleton pregnancies during the first trimester from October 2010 to September 2013. The demographic, work shift, and sleep data used in this analysis were collected from both in-person interviews and take-home questionnaires during the first visit, which took place between 6^0^ weeks and 13^6^ weeks gestation(24). Data used in this analysis was accessed from the National Institutes of Child Health and Development Data and Specimen Hub (DASH). All study participants provided written informed consent and study protocols were in accordance with the requirements of the respective Institutional Review Boards. The present analysis included participants who reported a current work shift and had preterm delivery, GDM, and preeclampsia outcome information available. Participants diagnosed with diabetes or chronic hypertension prior to pregnancy were excluded from the analyses.

### Demographic and health outcome data

For shift work exposure information, pregnant participants with current employment were grouped according to self-reported shift-work category. As afternoon and night shifts include working hours in the evening, these groups were combined to make an “evening shift” category; likewise, participants with irregular/on-call shifts or rotating shifts were combined to make an “irregular/rotating shift” category. Due to small numbers, participants who reported working a split shift (which features two or more shifts throughout the day) were excluded from the analysis. Pregnancy outcome information was ascertained by medical record abstraction and/or post-delivery questionnaire (when chart information was unavailable) and coded as binary variables as detailed before (24). Preterm birth was defined as birth prior to 37 weeks and 0 days from the estimated gestational age. Preeclampsia was determined using the adapted ACOG 2013 guidelines(25) and coded as positive if the participant was diagnosed with “eclampsia”, “severe preeclampsia”, or “mild preeclampsia” during their pregnancy. GDM was determined based on chart abstraction and glucose tolerance testing results, as previously described(22).

The following variables were included in the model adjusted for sociodemographic characteristics: age (continuous, years), pre-pregnancy smoking (dichotomous, “yes” if smoked tobacco in the 3 months prior to pregnancy), self-reported race and ethnicity of investigator-specified groups (Asian, Hispanic, Non-Hispanic Black or African-American, Non-Hispanic White, or Other), and adversity (ordinal variable that combines indicators for poverty, education, insurance, and partner status). Race and ethnicity are social constructs and the inclusion of a race and ethnicity covariate in this analysis is to reflect social experiences and bias relevant to adverse pregnancy outcomes. The adversity variable is a cumulative adversity score derived from federal poverty level (100-200% or <100%, +1), education (high school or less, +1), insurance (government/military/other, +1), and participant partner status (single, +1) to adjust for variables related to socio-economic adversity(26). Sensitivity analyses adjusted for change in BMI during pregnancy, derived as the difference between BMI at visit 1 and BMI at visit 3 (calculated as weight in kg / m^2^).

### Actigraphy substudy analysis

To evaluate sleep characteristics associated with shift-work schedule, actigraphy data from the Sleep Patterns and Quality Substudy were examined. This substudy enrolled 901 participants, of whom 782 had valid actigraphy data, between June 2011 and April 2013. Actigraphy measures of sleep were collected using a wrist-worn Actiwatch Spectrum (Philips Respironics) for 7 consecutive days during the second trimester (16^0^-23^6^ weeks gestation), as previously described(27). This substudy excluded participants younger than 18 years and individuals with pre-existing hypertension and/or pre-existing diabetes(27). The Actiwatch contains an accelerometer to distinguish sleep wake epochs and has been validated against the gold-standard measurement of polysomnography(28). For this analysis, we included participants with least 5 days of valid actigraphy data, which were assessed and scored by trained specialists at the Northwestern University reading center, as previously described(27). For the secondary analyses, sleep midpoint was calculated as the midpoint between sleep start time and wake time for the sleep episode; sleep timing variability was derived as the average standard deviation of sleep midpoint across all days with valid actigraphy data.

### Statistical analysis

Demographic and medical characteristics of the study population were characterized in total and stratified by shift work group. Distribution of categorical and continuous variables across work groups was assessed with Chi-square tests, Fisher’s exact test for categorical variables with cells ≤ 5, and 1-way analysis of variance (ANOVA) tests and considered significant at p<0.05. We fitted an unadjusted and two adjusted logistic regression models for each adverse pregnancy outcome of interest to test the association with shift work. For each outcome, to avoid adjusting for potential variables on the causal chain, the first adjusted models included only sociodemographic characteristics of adversity (ordinal), pre-pregnancy smoking (binary), self-reported race/ethnicity (categorical), and age (continuous); the second adjusted model included all of these covariates in addition to self-reported sleep duration category (categorical) and BMI (continuous) as covariates. Sensitivity analyses adjusted for difference in BMI between visit 1 and visit 3 (continuous).

### Mediation Analysis

We explored possible mediation of the relationship between shiftwork and adverse pregnancy outcomes by night-to-night variability in sleep timing, as measured by the standard deviation of sleep midpoint. Using the actigraphy data subset, we created logistic regression and linear models to measure the association between evening shift work (exposure), significantly associated adverse pregnancy outcome (outcome), and variability in sleep timing (mediator). Mediation analysis was conducted only if the proposed mediator was associated (p<0.05) with both the exposure and outcome. Mediation by variability in sleep timing was estimated using the “mediation” package(29) to calculate the average direct effect (ADE), the average causal mediation effect (ACME), the total effect, and the proportion of mediation. Uncertainty estimates were calculated with n=10,000 simulations using the quasi-Bayesian Monte Carlo method(30). Results with p<0.05 were considered statistically significant. Analyses were conducted in R version 4.1.1.

## RESULTS

### Sample characteristics

Of the enrolled nuMoM2b participants, 9,289 consented to their data being shared and maintained by NIH in the DASH database. Of these, a total of 5,192 participants without prior diabetes or chronic hypertension, who worked outside of the home, reported work shift history, and had pregnancy outcome data were included in the final analysis, as detailed in **Figure 1**. Comparing demographic characteristics of the participants who were (N=5,192) and were not included (N=4,097) in the analysis, included participants had lower BMI, were older, were more likely to identify as White race/ethnicity, report not smoking prior to pregnancy, have higher income, and have higher educational attainment. Additionally, included participants had a lower adversity score, less frequently self-reported long (>9 hours) sleep duration, and lower frequencies of all adverse pregnancy outcomes of interest (**Table 1**).

**Figure 1.**
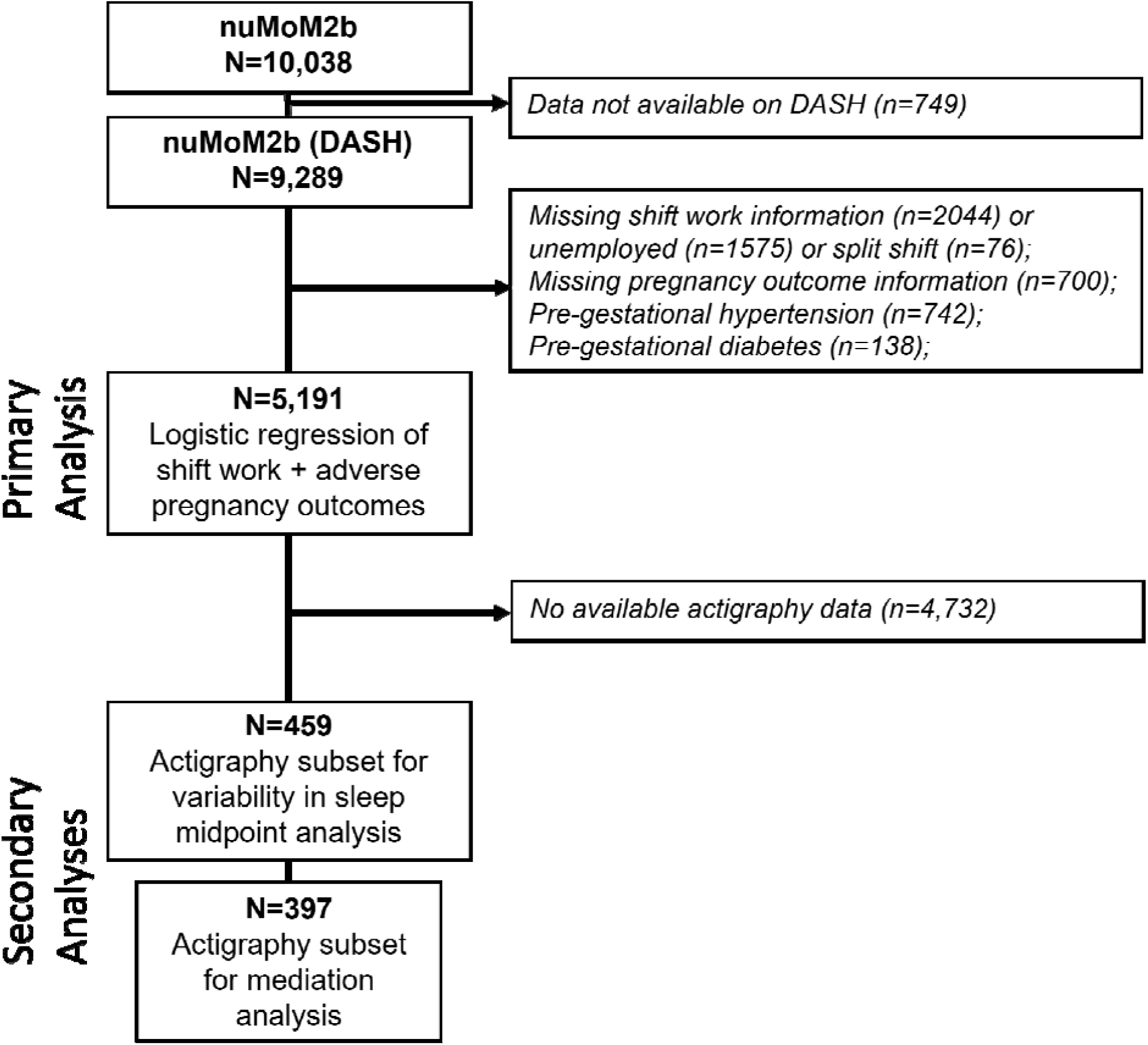
Flow chart depicting sample sizes of participants included in the primary and secondary analyses.

**Table 1.** Demographic and health characteristics of the analytical sample (n=5,192) from the nuMoM2b study.

|  | **All participants**<br>**(N=5,191)** | **Day**<br>**(N=3,833,**<br>**73.8%)** | **Evening**<br>**(N=616,**<br>**11.9%)** | **Irregular/**<br>**Rotating**<br>**(N=743,**<br>**14.3%)** | **p-value** |
| --- | --- | --- | --- | --- | --- |
| **\*BMI (mean (SD))** | 25.90 (5.77) | 25.85 (5.66) | 26.47 (6.36) | 25.70 (5.81) | 0.03 |
| **\*Age, years (mean (SD))** | 28.35 (5.07) | 29.18 (4.78) | 25.30 (5.06) | 26.62 (5.10) | <0.001 |
| **\*Pre-pregnancy smoking (Yes %)** | 735 (14.2) | 435 (11.4) | 139 (22.6) | 161 (21.7) | <0.001 |
| **\*Race/Ethnicity (%)** |  |  |  |  | <0.001 |
| *Asian* | 210 (4.0) | 166 (4.3) | 20 (3.2) | 24 (3.2) |  |
| *Hispanic* | 660 (12.7) | 441 (11.5) | 113 (18.3) | 106 (14.3) |  |
| *Non-Hispanic Black or African-American* | 405 (7.8) | 235 (6.1) | 110 (17.9) | 60 (8.1) |  |
| *Non-Hispanic White* | 3698 (71.2) | 2844 (74.2) | 340 (55.2) | 514 (69.2) |  |
| *Other* | 218 (4.2) | 146 (3.8) | 33 (5.4) | 39 (5.2) |  |
| **Income as % federal poverty threshold (mean (SD))** | 495.70 (299.74) | 535.69 (294.17) | 300.81 (244.47) | 413.54 (295.21) | <0.001 |
| **Income category as % federal poverty threshold** |  |  |  |  | <0.001 |
| *>200%* | 3663 (79.8) | 3005 (85.4) | 245 (52.6) | 413 (68.0) |  |
| *100-200%* | 555 (12.1) | 324 (9.2) | 125 (26.8) | 106 (17.5) |  |
| *<100%* | 373 (8.1) | 189 (5.4) | 96 (20.6) | 88 (14.5) |  |
| **Currently in school (Yes%)** | 1010 (19.5) | 637 (16.6) | 197 (32.0) | 176 (23.7) | <0.001 |
| **Education (%)** |  |  |  |  | <0.001 |
| *High school or less* | 514 (9.9) | 279 (7.3) | 131 (21.3) | 104 (14.0) |  |
| *Some college / Associate's* | 1382 (26.6) | 830 (21.7) | 294 (47.7) | 258 (34.7) |  |
| *College or more* | 3294 (63.5) | 2722 (71.1) | 191 (31.0) | 381 (51.3) |  |
| **Has a partner (Yes%)** | 5026 (96.9) | 3758 (98.1) | 560 (91.1) | 708 (95.4) | <0.001 |
| **Insurance (Private%)** |  | 3420 (89.5) | 393 (63.9) | 553 (74.9) | <0.001 |
| **\*Adversity (%)** |  |  |  |  | <0.001 |
| *0* | 3644 (70.2) | 2961 (77.3) | 259 (42.0) | 424 (57.1) |  |
| *1* | 893 (17.2) | 569 (14.8) | 156 (25.3) | 168 (22.6) |  |
| *2* | 465 (9.0) | 218 (5.7) | 139 (22.6) | 108 (14.5) |  |
| *≥3* | 189 (3.6) | 84 (2.2) | 62 (10.1) | 43 (5.8) |  |
| **\*Self-reported average sleep duration (%)** |  |  |  |  | <0.001 |
| *Short (<7 hours)* | 774 (15.0) | 499 (13.1) | 123 (20.0) | 152 (20.7) |  |
| *Average (7-9 hours)* | 3574 (69.2) | 2785 (73.0) | 320 (52.0) | 469 (63.7) |  |
| *Long (>9 hours)* | 817 (15.8) | 530 (13.9) | 172 (28.0) | 115 (15.6) |  |
| **Preeclampsia (Yes%)** | 394 (7.6) | 274 (7.1) | 59 (9.6) | 61 (8.2) | 0.084 |
| **Preterm birth (Yes%)** | 342 (6.6) | 237 (6.2) | 45 (7.3) | 60 (8.1) | 0.122 |
| **Gestational diabetes (Yes%)** | 203 (3.9) | 144 (3.8) | 33 (5.4) | 26 (3.5) | 0.135 |
| *Variables with an asterisk (\*) were included in the final fully adjusted model.* |  |  |  |  |  |

Of participants included in the analysis, approximately 73.8% reported working a day shift, 11.9% reported working an evening shift, and 14.3% reported working an irregular/rotating shift (**Supplemental Table 1**). Overall, demographic and health characteristics differed by shift work category, with evening and irregular/rotating shift workers being younger, more likely to smoke pre-pregnancy, identify as Black/African-American or Hispanic, have lower income, a current student, and/or have lower educational attainment, and be more likely to have both short (<7 hours) or long (>9 hours) self-reported sleep duration compared to day shift workers. Approximately 7.6% of individuals had a diagnosis of preeclampsia, 6.6% had preterm birth (<37 weeks), and 3.9% of pregnancies had a diagnosis of GDM.

### Associations between shift work and adverse pregnancy outcomes

The associations of shift work categories with pregnancy outcomes are shown in **Table 2**. Compared to day workers, those who worked evening shifts had 38% higher odds of developing preeclampsia in unadjusted analyses (unadjusted OR=1.38, 95% CI:1.02-1.84), but the association was attenuated and no longer significant after covariate adjustments (adjusted model 1 OR=1.25, 95% CI:0.91-1.71; adjusted model 2 OR=1.21, 95% CI:0.87-1.65).

**Table 2.** Associations between shift work categories and adverse pregnancy outcomes.

|  |  |  |  |
| --- | --- | --- | --- |
| **Shift Work Categories** | **Crude OR Estimate (95% CI)** | **Adjusted Model 1 OR\* Estimate (95% CI)** | **Adjusted Model 2 OR\* Estimate (95% CI)** |
| ***Preeclampsia*** |  |  |  |
| Day (ref) | 1.00 | 1.00 | 1.00 |
| Evening | **1.38 (1.02-1.84)** | 1.25 (0.91-1.71) | 1.21 (0.87-1.65) |
| Irregular/rotating | 1.16 (0.86-1.54) | 1.16 (0.85-1.55) | 1.17 (0.86-1.57) |
| ***Preterm Birth*** |  |  |  |
| Day (ref) | 1.00 | 1.00 | 1.00 |
| Evening | 1.20 (0.85-1.65) | 1.12 (0.78-1.58) | 1.10 (0.77-1.56) |
| Irregular/rotating | 1.33 (0.98-1.78) | 1.32 (0.97-1.78) | 1.25 (0.91-1.69) |
| ***Gestational Diabetes*** |  |  |  |
| Day (ref) | 1.00 | 1.00 | 1.00 |
| Evening | 1.45 (0.97-2.11) | **1.73 (1.12-2.60)** | **1.75 (1.12-2.66)** |
| Irregular/rotating | 0.93 (0.59-1.40) | 1.08 (0.68-1.64) | 1.07 (0.67-1.66) |
| Adjusted model 1 included adversity, pre-pregnancy smoking, race/ethnicity, and age as covariates; adjusted model 2 contained the same covariates as model 1 in addition to self-reported sleep duration and BMI. Analysis included n=3,833 day shift, n=616 evening shift, n=743 irregular/rotating shift workers. Estimates with p-values <0.05 are shown in bold. |  |  |  |

Comparatively, evening shift work was associated with 45% increased odds of GDM in unadjusted analyses which was not statistically significant (unadjusted OR=1.45, 95% CI:0.97-2.11), but this estimate increased and was significant after covariate adjustments (adjusted model 1 OR=1.73, 95% CI:1.12-2.60; adjusted model 2 OR=1.75, 95% CI:1.12-2.66). Unlike evening shift workers, pregnant participants who worked irregular or rotating shifts did not have higher odds of any the outcomes studied.

To check the consistency of associations across evening shift categories, we also performed a sensitivity analysis to compare results between working the afternoon shift or the night shift. Adjusted odds ratios were not significant for either category and preterm birth (afternoon OR=1.26, 95% CI:0.80-1.90; night OR=0.92, 95% CI:0.53-1.51), but afternoon shift was significantly associated with increased odds for preeclampsia (afternoon OR=1.54, 95% CI: 1.05-2.21; night OR=0.81, 95% CI:0.47-1.32) and night shift was significantly associated with increased odds for GDM (afternoon OR=1.50, 95% CI:0.82-2.58; night OR=2.06, 95% CI:1.14-3.52), suggesting that the association between evening shift and GDM was primarily driven by night shift workers.

### Secondary analysis of actigraphy data

Of those included in the primary analysis of shift work and adverse pregnancy outcomes, actigraphy data from the Sleep Patterns and Quality Substudy were available for 459 participants (**Supplemental Table 2**). Shift workers may have greater misalignment in night-to-night sleep timing than day workers due to differences between work hour schedule and social schedule or behavioral activity preference(31). Analysis of actigraphy measures during pregnancy supports this: on average, day workers have approximately 52 minutes variability in sleep timing, whereas sleep timing in evening and irregular/rotating shift workers varies by about 94 and 74 minutes, respectively (**Table 3**); in the sensitivity analysis evaluating differences by evening shift subcategory, night shift workers had a sleep timing variability of approximately 159 minutes, compared to 54 minutes for afternoon workers.

**Table 3.** Average sleep timing variability (minutes) by work shift category in secondary analysis of actigraphy data.

|  |  |  |  |  |  |  |
| --- | --- | --- | --- | --- | --- | --- |
| **Coefficients** | **Crude Estimate (Std Dev) of Sleep Timing Variability** | **t-value (crude)** | **p-value (crude)** | **Adjusted Estimate (Std Dev) of Sleep Timing Variability** | **t-value (adjusted)** | **p-value (adjusted)** |
| Intercept | 51.889 (2.99) | 17.37 | **<2e-16** | 53.551 (20.22) | 2.65 | **0.008** |
| Evening (n=65) | 42.537 (7.39) | 5.76 | **1.55e-08** | 47.362 (7.95) | 5.96 | **5.36e-09** |
| Irregular/rotating (n=62) | 21.662 (7.53) | 2.88 | **0.004** | 24.253 (7.96) | 3.05 | **0.003** |
| Note: Participants with chronic hypertension or diabetes prior to pregnancy were excluded from the analysis, for a total of 459 included participants (ref=day shift, n=332). Adjusted models included adversity, pre-pregnancy smoking, race/ethnicity, self-reported sleep duration category, age, and BMI as covariates. P-values <0.05 are shown in bold. |  |  |  |  |  |  |

### Mediation analysis with actigraphy data

Next, we conducted a mediation analysis between evening shift work, variability in sleep timing, and GDM. In unadjusted analyses, shift work was associated with both GDM and sleep timing variability. Likewise, when sleep timing variability was included as a model predicting GDM, the crude odds ratio decreased from 1.82 to 1.47 and shift work was no longer associated with GDM. These findings suggest mediation of the relationship between shift work and GDM by sleep timing variability. Therefore, we conducted an exploratory mediation analysis comparing day and evening shift workers with actigraphy data, for a total of 397 participants with 18 total cases of GDM. The average direct effect (ADE) of evening shift work on GDM was not significant (ADE=0.05, 95% CI:-0.015-0.14, p=0.152) after adjusting for sleep timing variability, but the indirect effect [the average causal mediation effect (ACME)] was statistically significant (ACME=0.018, 95% CI:0.004-0.04, p<0.01); the proportion mediated was 0.27 (0.018/0.067), suggesting 27% of the association between shift work and GDM was mediated by sleep timing variability (**Figure 2**).

**Figure 2.**
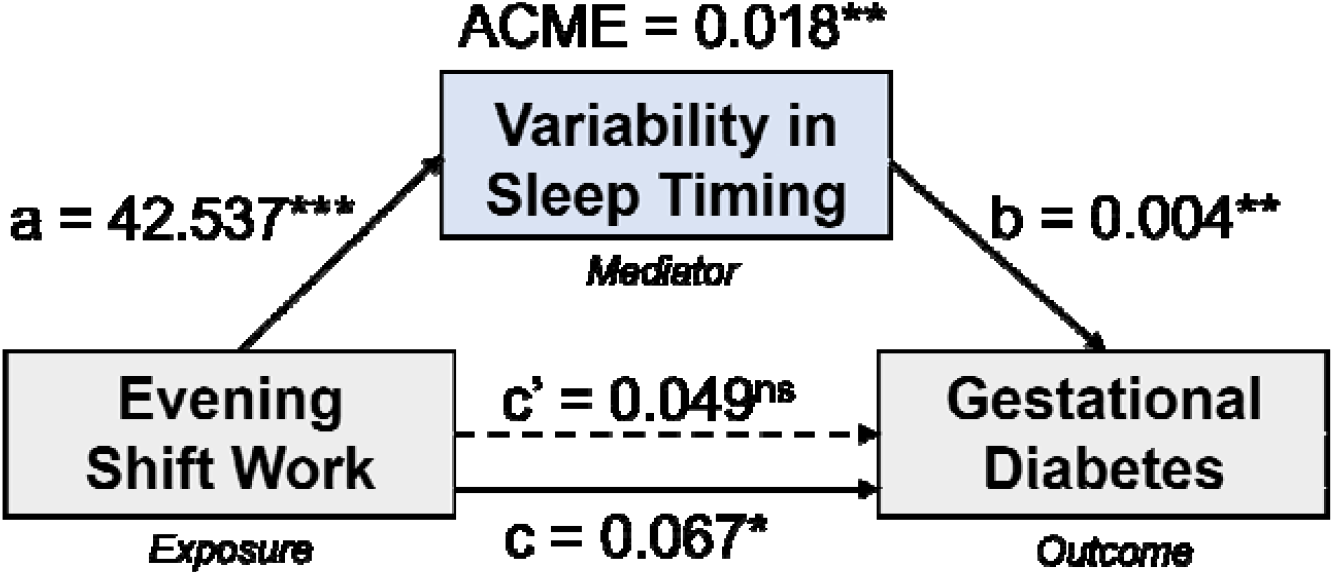
Diagram of mediation analysis results examining whether association between evening shift work (binary exposure) and GDM (binary outcome) is mediated by variability in objective sleep timing (continuous, minutes). Day workers (n=332) and evening workers (n=65) with actigraphy data were compared. Regression coefficients are presented, with path a representing the effect of evening shift work on sleep timing variability, path b representing the effect of variability in sleep timing on GDM, path c’ representing the direct effect of evening shift work on GDM (the average direct effect, ADE), the average causal mediation effect (ACME) representing the direct effect of variability in sleep timing on GDM, and path c representing the total effect of evening shift work on GDM when sleep timing variability is included in the model. Asterisks represent *p<0.05, **p<0.01, ***p<0.001, ns = not significant.

## DISCUSSION

In this study we evaluated the association between shift work and cardiometabolic disease during pregnancy in a prospective birth cohort study. We show that evening (afternoon or night) shift work during pregnancy is associated with higher risk of GDM. An exploratory mediation analysis suggested that the relationship between evening shift work and GDM may be mediated by variability in sleep timing. Compared to day workers, evening shift workers had the greatest irregularity in sleep timing, with approximately 43 minutes greater sleep timing variability. Overall, our results support an association between evening shift work, sleep timing variability, and GDM during pregnancy in nulliparous participants in a large, prospective birth cohort study.

There are a number of mechanisms that may link evening shift work to adverse pregnancy outcomes, particularly GDM. For example, circadian disruption due to shift work may lead to inflammation and immune dysregulation, which are believed to play key roles in the development of both preeclampsia(32) and GDM(33). Likewise, light exposure out of sync with the endogenous circadian system can suppress production of melatonin, a hormone which acts as an antioxidant and plays a role in glucose regulation(34). Altered levels of cortisol could also lead to impaired glucose tolerance and GDM, as cortisol acts as an insulin antagonist and may mediate the associations between shift work and cardiometabolic disease risk(35). For example, night shift workers produce higher levels of cortisol compared to day workers(36,37), and this effect is more pronounced in younger (<40 years old) workers(38). Since the majority of pregnancies occur before age 40 and shift workers tend to be younger, the greater exposure among younger individuals to the impacts of shift work on cortisol levels may be especially relevant for pregnancy and GDM. Few animal studies of circadian misalignment during pregnancy and gestational glucose tolerance exist, but a simulated shift work schedule during pregnancy caused impaired glucose tolerance during early (but not late) pregnancy in sheep(39). Variability in eating times due to shift work may also alter cortisol levels and cause glucose intolerance and insulin resistance; in a clinical trial, late eating prior to sleep resulted in higher post-meal glycemia and cortisol(40). Similarly, short sleep and disrupted sleep may promote appetite dysregulation by altering hunger and satiety hormones(41).

During pregnancy, increased metabolic demands and resulting physiological changes may serve as a “stress test” to reveal early forms of disease(42). Pregnant people who develop GDM that resolves following delivery are at an increased risk of later developing type 2 diabetes(43,44), and offspring of individuals who had GDM have increased risk of developing chronic cardiometabolic disease(45–47). Thus, prevention or treatment of GDM may reduce morbidity in both mothers and offspring. While shift work is a well-established risk factor for developing T2D(3,4,48), our findings support the need to further investigate whether shift work prior to or during pregnancy is a risk factor for GDM, and to identify potential mitigation strategies in those at greatest risk for adverse pregnancy outcomes.

Sleep timing has previously been shown to be an important factor for adverse pregnancy outcomes among individuals in the nuMoM2b cohort, with self-reported late midpoint of sleep (>5 AM) associated with preterm birth(21) and GDM(22), and objectively assessed (by actigraphy) later sleep midpoint (>5 AM) also associated with GDM(23). Our results build on these prior findings by evaluating the contributions of shift work and variability in objective sleep timing. Irregular sleep schedules may be a common driver of circadian misalignment in the general population, but shift workers likely have even greater variability in sleep schedules due to more extreme differences in activity timing on work and non-work days. For example, in the Hispanic Community Health Study/ Study of Latinos sleep ancillary study “Sueño”, numerous actigraphy measures differ between shift and day workers, with later objective sleep midpoint and greater variation in sleep timing among participants working night or irregular shifts(31), similar to our findings in pregnant participants.

Our results are not entirely consistent with the overall evidence between shift work and adverse pregnancy outcomes. While our analysis did not support an association with preterm birth, rotating shift and night shift work has previously been linked to approximately 13% and 21% increased odds of preterm birth in systematic reviews and meta-analyses(10,49). Additionally, our results differ from those of the one prior study of night shift work and GDM; this prospective birth cohort study in Japan reported no increased odds of GDM among fixed night shift workers(13). Additionally, although prior studies report associations between rotating shifts (50,51) and night shifts and preeclampsia(51–53), our adjusted results do not support an association between evening shift work and preeclampsia. There are a number of factors which could contribute to the heterogeneity in results; in particular, many of the prior cohorts were not based in the U.S. Some of the discrepancies in findings may be related to different social-ecological factors linked to shift work during pregnancy. For example, heterogeneity in healthcare systems, access to prenatal care, and policies affecting whether someone is able to take an occupational leave during pregnancy may modify the associations between shift work and health outcomes between countries. Additionally, smaller sample sizes, enrollment of participants later in pregnancy or post-pregnancy, differences in parity, adjustment for confounders, and differences in exposure measurement and shift work categorization in prior studies also could have contributed to between-study differences in effect estimates.

This study has a number of strengths and limitations which should be taken into consideration when interpreting the results. Because of the prospective nature of this cohort and enrollment of nulliparous participants during early pregnancy, the design of nuMoM2b is well-suited to capture adverse pregnancy outcomes. However, information on shift work intensity and duration was unavailable, limiting the ability to evaluate dose-response relationships; this information would also have been useful to evaluate whether the lower sleep timing variability in irregular/rotating shift workers compared to evening shift workers was due to shift rotation schedules and/or intensity of rotations. Information on meal timing and chronotype, other possible links between shift work and GDM, were also unavailable. However, adjustment for BMI or for change in BMI from third to first trimester did not decrease the strength of association between evening shift work and GDM, in line with reports of cardiometabolic effects of shift work independent of adiposity and BMI(7), suggesting health effects of evening shift work separate from BMI. While shift work status was ascertained during the first trimester, the actigraphy substudy took place during the second trimester; therefore, it is possible that some of the participants who reported a work shift at the beginning of pregnancy left their job position prior to actigraphy measurement, leading to misclassification of exposure. Additionally, participants included in the analysis had higher overall indicators of socioeconomic status and lower prevalence of adverse pregnancy outcomes compared to those not included, suggesting possible limitation of the generalizability of results. Our exploratory mediation analysis was conducted in a small sample, and results should be interpreted with caution. Additionally, the overall occurrence of GDM in this nulliparous cohort was approximately 3.9%, lower than the 2016 U.S. nulliparous prevalence of 5.2%(54); while this difference may be due to the exclusion criteria and eligibility restrictions (nulliparous), the incidence of GDM is increasing and more research is needed to better understand the role of sleep timing in disease pathology. Sleep schedules are potentially modifiable, and these findings support further investigation of sleep regularity as a possible area for intervention and health promotion during pregnancy. However, modifying sleep schedules may be more difficult in the context of shiftwork, requiring a consistent later sleep schedule or leaving the non-day shift; in some countries, pregnant people are prohibited from working night shifts(8), which may be viewed as controversial and infringing on personal freedoms. Therefore, developing guidelines around shiftwork during pregnancy are complicated, and a number of factors, such as those recently outlined here(55), must be taken into account in addressing related health outcomes.

## CONCLUSION

In conclusion, evening shift work among pregnant people is associated with increased odds of developing GDM. When actigraphy measures were compared between different shift work categories, pregnant participants who reported working evening shifts or irregular/rotating shifts had greater variability in sleep timing compared to day workers. While we are somewhat limited by small group sizes in the actigraphy analyses, these findings implicate objectively-measured sleep timing variability in the causal path between shift work and GDM. Overall, our results support the need to further consider sleep and behavior rhythms in cardiometabolic disease during pregnancy. Sleep schedules are modifiable and may offer an area for intervention to improve pregnancy outcomes, particularly in people with non-day work shifts. Future studies should investigate whether consistent sleep schedules and decreased variability in sleep timing may improve cardiometabolic outcomes during pregnancy.

## Data Availability

All data analyzed in the present work are hosted on the NICHD Data Specimen Hub and are available with proper institutional application and approval.

## ACKNOWLEDGEMENTS

Supported by funding from the National Institutes of Health (NIH-NHLBI T32HL007901 [to DW], and R35 HL135818 [to SR]). We acknowledge NICHD DASH for providing the Nulliparous Pregnancy Outcomes Study: Monitoring Mothers-to-be data that was used for this research. This study was supported by grant funding from the Eunice Kennedy Shriver National Institute of Child Health and Human Development (NICHD) and the National Heart, Lung, and Blood Institute (NHLBI): U10 HD063036; U10 HD063072; U10 HD063047; U10 HD063037; U10 HD063041; U10 HD063020; U10 HD063046; U10 HD063048; U10 HD063053; and NHLBI R01 HL105549. In addition, support was provided by Clinical and Translational Science Institutes: UL1TR001108 and UL1TR000153.

---

**Supplemental Table 1.** Demographic and health characteristics of the available nuMoM2b DASH dataset.

|  | **All**<br>**(N=9,289)** | **Excluded**<br>**(N=4,097)** | **Included**<br>**(N=5,191)** | **p-value** |
| --- | --- | --- | --- | --- |
| **BMI (mean (SD))** | 26.37 (6.34) | 26.99<br>(6.97) | 25.90 (5.77) | <0.001 |
| **Age, years (mean (SD))** | 26.91 (5.68) | 25.08<br>(5.89) | 28.35 (5.07) | <0.001 |
| **Pre-pregnancy smoking (N(Yes%))** | 1671 (18.0) | 936 (22.9) | 735 (14.2) | <0.001 |
| **Self-reported race/ethnicity (N(%))** |  |  |  | <0.001 |
| *Asian* | 365 (3.9) | 155 (3.8) | 210 (4.0) |  |
| *Hispanic* | 1599 (17.2) | 939 (23.0) | 660 (12.7) |  |
| *Non-Hispanic Black or African-American* | 1248 (13.4) | 843 (20.6) | 404 (7.8) |  |
| *Non-Hispanic White* | 5595 (60.3) | 1897 (46.4) | 3698 (71.2) |  |
| *Other* | 472 (5.1) | 254 (6.2) | 218 (4.2) |  |
| **Income as % federal poverty threshold (mean (SD))** | 431.39<br>(309.7) | 331.19<br>(298.1) | 495.70<br>(299.7) | <0.001 |
| **Income category as % federal poverty threshold (N(%))** |  |  |  | <0.001 |
| *>200%* | 5227 (69.3) | 1564 (53.1) | 3663 (79.8) |  |
| *100-200%* | 1094 (14.5) | 539 (18.3) | 555 (12.1) |  |
| *<100%* | 1217 (16.1) | 844 (28.6) | 373 (8.1) |  |
| **Currently in school (Yes%)** | 2258 (24.3) | 1248 (30.6) | 1010 (19.5) |  |
| **Education (N(%))** |  |  |  | <0.001 |
| *High school or less* | 1850 (20.0) | 1336 (32.7) | 514 (9.9) |  |
| *Some college / Associate's* | 2764 (29.8) | 1382 (33.9) | 1381 (26.6) |  |
| *College or more* | 4657 (50.2) | 1363 (33.4) | 3294 (63.5) |  |
| **Has a partner (N(Yes%))** | 8738 (94.3) | 3712 (91.0) | 5025 (96.9) | <0.001 |
| **Health insurance (N(Private%))** | 6510 (70.6) | 2144 (53.1) | 4366 (84.4) | <0.001 |
| **Cumulative adversity score (N(%))** |  |  |  | <0.001 |
| *0* | 5142 (55.4) | 1498 (36.6) | 3644 (70.2) |  |
| *1* | 1796 (19.4) | 903 (22.1) | 892 (17.2) |  |
| *2* | 1530 (16.5) | 1065 (26.1) | 465 (9.0) |  |
| *≥3* | 811 (8.7) | 622 (15.2) | 189 (3.6) |  |
| **Self-reported average sleep duration (N(%))** |  |  |  | <0.001 |
| *Short (<7 hours)* | 1071 (15.0) | 297 (15.0) | 774 (15.0) |  |
| *Average (7-9 hours)* | 4575 (64.0) | 1001 (50.4) | 3574 (69.2) |  |
| *Long (>9 hours)* | 1504 (21.0) | 687 (34.6) | 817 (15.8) |  |
| **Preeclampsia (N(Yes%))** | 780 (8.9) | 386 (10.8) | 393 (7.6) | <0.001 |
| **Preterm birth (N(Yes%))** | 725 (8.2) | 383 (10.6) | 341 (6.6) | <0.001 |
| **Gestational diabetes (N(Yes%))** | 376 (4.3) | 173 (4.9) | 203 (3.9) | 0.029 |

**Supplemental Table 2.** Demographic and health characteristics of the nuMoM2b participants included in the sleep substudy analysis.

|  | **All**<br>**(N=459)** | **Day**<br>**(N=332,**<br>**72.3%)** | **Evening**<br>**(N=65,**<br>**14.2%)** | **Irregular/**<br>**Rotating**<br>**(N=62,**<br>**13.5%)** | **p-**<br>**value** |
| --- | --- | --- | --- | --- | --- |
| **\*BMI (mean (SD))** | 25.95 (5.89) | 25.91 (5.79) | 27.02 (6.51) | 25.06 (5.66) | 0.167 |
| **\*Age, years (mean (SD))** | 27.86 (5.18) | 28.87 (4.99) | 24.97 (5.11) | 25.45 (4.30) | <0.001 |
| **\*Pre-pregnancy smoking**<br>**(Yes %)** | 57 (12.4) | 34 (10.2) | 14 (21.5) | 9 (14.5) | 0.036 |
| **Self-reported race/ethnicity**<br>**(N(%))** |  |  |  |  | 0.046 |
| Asian | 15 (3.3) | 11 (3.3) | 3 (4.6) | 1 (1.6) |  |
| Hispanic | 48 (10.5) | 26 (7.8) | 14 (21.5) | 8 (12.9) |  |
| Non-Hispanic Black or African-American | 41 (8.9) | 29 (8.7) | 8 (12.3) | 4 (6.5) |  |
| Non-Hispanic White | 330 (71.9) | 249 (75.0) | 37 (56.9) | 44 (71.0) |  |
| Other | 25 (5.4) | 17 (5.1) | 3 (4.6) | 5 (8.1) |  |
| **Income as % federal poverty**<br>**threshold (mean (SD))** | 423.44<br>(272.81) | 462.65<br>(273.27) | 283.58<br>(219.14) | 312.51<br>(244.26) | <0.001 |
| **Income category as %**<br>**federal poverty threshold**<br>**(N(%))** |  |  |  |  | <0.001 |
| >200% | 299 (75.3) | 249 (82.5) | 23 (51.1) | 27 (54.0) |  |
| 100-200% | 61 (15.4) | 33 (10.9) | 14 (31.1) | 14 (28.0) |  |
| <100% | 37 (9.3) | 20 (6.6) | 8 (17.8) | 9 (18.0) |  |
| **Currently in school (Yes%)** | 93 (20.3) | 61 (18.4) | 18 (27.7) | 14 (22.6) | 0.206 |
| **Education (N(%))** |  |  |  |  | <0.001 |
| High school or less | 42 (9.2) | 22 (6.6) | 13 (20.0) | 7 (11.3) |  |
| Some college / Associate's | 168 (36.6) | 98 (29.5) | 38 (58.5) | 32 (51.6) |  |
| College or more | 249 (54.2) | 212 (63.9) | 14 (21.5) | 23 (37.1) |  |
| **Has a partner (Yes%)** | 444 (96.7) | 328 (98.8) | 59 (90.8) | 57 (91.9) | <0.001 |
| **Insurance (Private%)** | 389 (84.9) | 298 (90.0) | 45 (69.2) | 46 (74.2) | <0.001 |
| **Cumulative adversity score**<br>**(N(%))** |  |  |  |  | <0.001 |
| 0 | 304 (66.2) | 252 (75.9) | 25 (38.5) | 27 (43.5) |  |
| 1 | 104 (22.7) | 57 (17.2) | 26 (40.0) | 21 (33.9) |  |
| 2 | 37 (8.1) | 15 (4.5) | 9 (13.8) | 13 (21.0) |  |
| ≥3 | 14 (3.1) | 8 (2.4) | 5 (7.7) | 1 (1.6) |  |
| **Self-reported average sleep**<br>**duration (N(%))** |  |  |  |  | 0.639 |
| Short (<7 hours) | 76 (16.6) | 55 (16.6) | 9 (14.1) | 12 (19.4) |  |
| Average (7-9 hours) | 309 (67.6) | 222 (67.1) | 43 (67.2) | 44 (71.0) |  |
| Long (>9 hours) | 72 (15.8) | 54 (16.3) | 12 (18.8) | 6 (9.7) |  |
| **Preeclampsia (Yes%)** | 32 (7.0) | 24 (7.2) | 5 (7.7) | 3 (4.8) | 0.771 |
| **Preterm birth (Yes%)** | 28 (6.1) | 21 (6.3) | 4 (6.2) | 3 (4.8) | 0.904 |
| **Gestational diabetes (Yes%)** | 19 (4.1) | 11 (3.3) | 7 (10.8) | 1 (1.6) | 0.012 |

## Bibliography

1. McMenamin T. A time to work: recent trends in shift work and flexible schedules. Mon Labor Rev [Internet]. 2007 Dec 1 [cited 2021 Nov 3]; Available from: https://www.bls.gov/opub/mlr/2007/12/art1full.pdf

2. U.S. Census Bureau. Maternity leave and employment patterns of first-time mothers: 1961–2008: current population reports P70-128 [Internet]. 2011 Oct [cited 2021 Oct 7]. Available from: https://www.census.gov/prod/2011pubs/p70-128.pdf

3. Vetter C, Dashti HS, Lane JM, Anderson SG, Schernhammer ES, Rutter MK, et al. Night shift work, genetic risk, and type 2 diabetes in the UK biobank. Diabetes Care. 2018 Apr;41(4):762–9.

4. Gan Y, Yang C, Tong X, Sun H, Cong Y, Yin X, et al. Shift work and diabetes mellitus: a meta-analysis of observational studies. Occup Environ Med. 2015 Jan;72(1):72–8.

5. Dutheil F, Baker JS, Mermillod M, De Cesare M, Vidal A, Moustafa F, et al. Shift work, and particularly permanent night shifts, promote dyslipidaemia: A systematic review and meta-analysis. Atherosclerosis. 2020 Nov;313:156–69.

6. Kecklund G, Axelsson J. Health consequences of shift work and insufficient sleep. BMJ. 2016 Nov 1;355:i5210.

7. Sweeney E, Yu ZM, Dummer TJB, Cui Y, DeClercq V, Forbes C, et al. The relationship between anthropometric measures and cardiometabolic health in shift work: findings from the Atlantic PATH Cohort Study. Int Arch Occup Environ Health. 2020 Jan;93(1):67–76.

8. IARC Working Group on the Identification of Carcinogenic Hazards to Humans. Night Shift Work. Lyon (FR): International Agency for Research on Cancer; 2020.

9. Leproult R, Van Cauter E. Role of sleep and sleep loss in hormonal release and metabolism. Endocr Dev. 2010;17:11–21.

10. Cai C, Vandermeer B, Khurana R, Nerenberg K, Featherstone R, Sebastianski M, et al. The impact of occupational shift work and working hours during pregnancy on health outcomes: a systematic review and meta-analysis. Am J Obstet Gynecol. 2019 Dec;221(6):563–76.

11. Specht IO, Hammer PEC, Flachs EM, Begtrup LM, Larsen AD, Hougaard KS, et al. Night work during pregnancy and preterm birth-A large register-based cohort study. PLoS ONE. 2019 Apr 18;14(4):e0215748.

12. Kader M, Bigert C, Andersson T, Selander J, Bodin T, Skröder H, et al. Shift and night work during pregnancy and preterm birth-a cohort study of Swedish health care employees. Int J Epidemiol. 2021 Jul 1;

13. Suzumori N, Ebara T, Matsuki T, Yamada Y, Kato S, Omori T, et al. Effects of long working hours and shift work during pregnancy on obstetric and perinatal outcomes: A large prospective cohort study-Japan Environment and Children’s Study. Birth. 2020 Mar;47(1):67–79.

14. Roenneberg T, Allebrandt KV, Merrow M, Vetter C. Social jetlag and obesity. Curr Biol. 2012 May 22;22(10):939–43.

15. Parsons MJ, Moffitt TE, Gregory AM, Goldman-Mellor S, Nolan PM, Poulton R, et al. Social jetlag, obesity and metabolic disorder: investigation in a cohort study. Int J Obes (Lond). 2015 May;39(5):842–8.

16. Wong PM, Hasler BP, Kamarck TW, Muldoon MF, Manuck SB. Social jetlag, chronotype, and cardiometabolic risk. J Clin Endocrinol Metab. 2015 Dec;100(12):4612–20.

17. Taylor BJ, Matthews KA, Hasler BP, Roecklein KA, Kline CE, Buysse DJ, et al. Bedtime variability and metabolic health in midlife women: the SWAN sleep study. Sleep. 2016 Feb 1;39(2):457–65.

18. Gooley JJ. How Much Day-To-Day Variability in Sleep Timing Is Unhealthy? Sleep. 2016 Feb 1;39(2):269–70.

19. Huang T, Redline S. Cross-sectional and Prospective Associations of Actigraphy-Assessed Sleep Regularity With Metabolic Abnormalities: The Multi-Ethnic Study of Atherosclerosis. Diabetes Care. 2019 Aug;42(8):1422–9.

20. Huang T, Mariani S, Redline S. Sleep Irregularity and Risk of Cardiovascular Events: The Multi-Ethnic Study of Atherosclerosis. J Am Coll Cardiol. 2020 Mar 10;75(9):991–9.

21. Facco FL, Parker CB, Hunter S, Reid KJ, Zee PP, Silver RM, et al. Later sleep timing is associated with an increased risk of preterm birth in nulliparous women. American Journal of Obstetrics & Gynecology MFM. 2019 Nov;1(4):100040.

22. Facco FL, Parker CB, Hunter S, Reid KJ, Zee PC, Silver RM, et al. Association of Adverse Pregnancy Outcomes With Self-Reported Measures of Sleep Duration and Timing in Women Who Are Nulliparous. J Clin Sleep Med. 2018 Dec 15;14(12):2047–56.

23. Facco FL, Grobman WA, Reid KJ, Parker CB, Hunter SM, Silver RM, et al. Objectively measured short sleep duration and later sleep midpoint in pregnancy are associated with a higher risk of gestational diabetes. Am J Obstet Gynecol. 2017 Oct;217(4):447.e1-447.e13.

24. Haas DM, Parker CB, Wing DA, Parry S, Grobman WA, Mercer BM, et al. A description of the methods of the Nulliparous Pregnancy Outcomes Study: monitoring mothers-to-be (nuMoM2b). Am J Obstet Gynecol. 2015 Apr;212(4):539.e1-539.e24.

25. The American College of O and G. Hypertension in pregnancy. Report of the American College of Obstetricians and Gynecologists’ Task Force on Hypertension in Pregnancy. Obstet Gynecol. 2013 Nov;122(5):1122–31.

26. Anand KJS, Rigdon J, Rovnaghi CR, Qin F, Tembulkar S, Bush N, et al. Measuring socioeconomic adversity in early life. Acta Paediatr. 2019 Jul;108(7):1267–77.

27. Reid KJ, Facco FL, Grobman WA, Parker CB, Herbas M, Hunter S, et al. Sleep During Pregnancy: The nuMoM2b Pregnancy and Sleep Duration and Continuity Study. Sleep. 2017 May 1;40(5).

28. Weiss AR, Johnson NL, Berger NA, Redline S. Validity of activity-based devices to estimate sleep. J Clin Sleep Med. 2010 Aug 15;6(4):336–42.

29. Tingley D, Yamamoto T, Hirose K, Keele L, Imai K. mediation:R package for causal mediation analysis. J Stat Softw. 2014;59(5).

30. Imai K, Keele L, Tingley D. A general approach to causal mediation analysis. Psychol Methods. 2010 Dec;15(4):309–34.

31. Reid KJ, Weng J, Ramos AR, Zee PC, Daviglus M, Mossavar-Rahmani Y, et al. Impact of shift work schedules on actigraphy-based measures of sleep in Hispanic workers: results from the Hispanic Community Health Study/Study of Latinos ancillary Sueño study. Sleep. 2018 Oct 1;41(10).

32. Harmon AC, Cornelius DC, Amaral LM, Faulkner JL, Cunningham MW, Wallace K, et al. The role of inflammation in the pathology of preeclampsia. Clin Sci. 2016 Mar;130(6):409–19.

33. Lowe LP, Metzger BE, Lowe WL, Dyer AR, McDade TW, McIntyre HD, et al. Inflammatory mediators and glucose in pregnancy: results from a subset of the Hyperglycemia and Adverse Pregnancy Outcome (HAPO) Study. J Clin Endocrinol Metab. 2010 Dec;95(12):5427–34.

34. Karamitri A, Jockers R. Melatonin in type 2 diabetes mellitus and obesity. Nat Rev Endocrinol. 2019 Feb;15(2):105–25.

35. Ritonja J, Aronson KJ, Day AG, Korsiak J, Tranmer J. Investigating cortisol production and pattern as mediators in the relationship between shift work and cardiometabolic risk. Can J Cardiol. 2018 May;34(5):683–9.

36. Zhang Y, Shen J, Zhou Z, Sang L, Zhuang X, Chu M, et al. Relationships among shift work, hair cortisol concentration and sleep disorders: a cross-sectional study in China. BMJ Open. 2020 Nov 14;10(11):e038786.

37. Li J, Bidlingmaier M, Petru R, Pedrosa Gil F, Loerbroks A, Angerer P. Impact of shift work on the diurnal cortisol rhythm: a one-year longitudinal study in junior physicians. J Occup Med Toxicol. 2018 Aug 14;13:23.

38. Manenschijn L, van Kruysbergen RGPM, de Jong FH, Koper JW, van Rossum EFC. Shift work at young age is associated with elevated long-term cortisol levels and body mass index. J Clin Endocrinol Metab. 2011 Nov;96(11):E1862–5.

39. Gatford KL, Kennaway DJ, Liu H, Kleemann DO, Kuchel TR, Varcoe TJ. Simulated shift work disrupts maternal circadian rhythms and metabolism, and increases gestation length in sheep. J Physiol (Lond). 2019 Apr;597(7):1889–904.

40. Gu C, Brereton N, Schweitzer A, Cotter M, Duan D, Børsheim E, et al. Metabolic Effects of Late Dinner in Healthy Volunteers-A Randomized Crossover Clinical Trial. J Clin Endocrinol Metab. 2020 Aug 1;105(8).

41. Reutrakul S, Van Cauter E. Sleep influences on obesity, insulin resistance, and risk of type 2 diabetes. Metab Clin Exp. 2018 Jul;84:56–66.

42. Williams D. Pregnancy: a stress test for life. Curr Opin Obstet Gynecol. 2003 Dec;15(6):465–71.

43. Daly B, Toulis KA, Thomas N, Gokhale K, Martin J, Webber J, et al. Increased risk of ischemic heart disease, hypertension, and type 2 diabetes in women with previous gestational diabetes mellitus, a target group in general practice for preventive interventions: A population-based cohort study. PLoS Med. 2018 Jan 16;15(1):e1002488.

44. Saravanan P, Diabetes in Pregnancy Working Group, Maternal Medicine Clinical Study Group, Royal College of Obstetricians and Gynaecologists, UK. Gestational diabetes: opportunities for improving maternal and child health. Lancet Diabetes Endocrinol. 2020 Sep;8(9):793–800.

45. Yu Y, Arah OA, Liew Z, Cnattingius S, Olsen J, Sørensen HT, et al. Maternal diabetes during pregnancy and early onset of cardiovascular disease in offspring: population based cohort study with 40 years of follow-up. BMJ. 2019 Dec 4;367:l6398.

46. Boney CM, Verma A, Tucker R, Vohr BR. Metabolic syndrome in childhood: association with birth weight, maternal obesity, and gestational diabetes mellitus. Pediatrics. 2005 Mar;115(3):e290–6.

47. Vääräsmäki M, Pouta A, Elliot P, Tapanainen P, Sovio U, Ruokonen A, et al. Adolescent manifestations of metabolic syndrome among children born to women with gestational diabetes in a general-population birth cohort. Am J Epidemiol. 2009 May 15;169(10):1209–15.

48. Gao Y, Gan T, Jiang L, Yu L, Tang D, Wang Y, et al. Association between shift work and risk of type 2 diabetes mellitus: a systematic review and dose-response meta-analysis of observational studies. Chronobiol Int. 2020 Jan;37(1):29–46.

49. Mozurkewich EL, Luke B, Avni M, Wolf FM. Working conditions and adverse pregnancy outcome: a meta-analysis. Obstet Gynecol. 2000 Apr;95(4):623–35.

50. Wergeland E, Strand K. Working conditions and prevalence of pre-eclampsia, Norway 1989. International Journal of Gynecology & Obstetrics. 1997 Aug;58(2):189–96.

51. Chang P-J, Chu L-C, Hsieh W-S, Chuang Y-L, Lin S-J, Chen P-C. Working hours and risk of gestational hypertension and pre-eclampsia. Occup Med (Lond). 2010 Jan;60(1):66–71.

52. Haelterman E, Marcoux S, Croteau A, Dramaix M. Population-based study on occupational risk factors for preeclampsia and gestational hypertension. Scand J Work Environ Health. 2007 Aug;33(4):304–17.

53. Nugteren JJ, Snijder CA, Hofman A, Jaddoe VWV, Steegers EAP, Burdorf A. Work-related maternal risk factors and the risk of pregnancy induced hypertension and preeclampsia during pregnancy. The Generation R Study. PLoS ONE. 2012 Jun 15;7(6):e39263.

54. Deputy NP, Kim SY, Conrey EJ, Bullard KM. Prevalence and Changes in Preexisting Diabetes and Gestational Diabetes Among Women Who Had a Live Birth - United States, 2012-2016. MMWR Morb Mortal Wkly Rep. 2018 Nov 2;67(43):1201–7.

55. Gurubhagavatula I, Barger LK, Barnes CM, Basner M, Boivin DB, Dawson D, et al. Guiding principles for determining work shift duration and addressing the effects of work shift duration on performance, safety, and health. Sleep. 2021 Jul 15;

